# Reduction in circulating monocytes correlates with persistent post-COVID pulmonary fibrosis in multi-omic comparison of long-haul COVID and IPF

**DOI:** 10.1101/2022.09.30.22280468

**Authors:** Grace C. Bingham, Lyndsey M. Muehling, Chaofan Li, Yong Huang, Daniel Abebayehu, Imre Noth, Jie Sun, Judith A. Woodfolk, Thomas H. Barker, Catherine Bonham

## Abstract

Rationale: Up to 30% of COVID-19 patients experience persistent sequelae, including dyspnea, restrictive physiology, and early radiographic signs of pulmonary fibrosis (PF). The mechanisms that provoke post-COVID progressive PF are poorly understood, and biomarkers to identify at-risk patients are urgently needed. Methods: We evaluated a cohort of 14 symptomatic COVID survivors with impaired respiratory function and imaging worrisome for developing PF, including bilateral reticulation, traction bronchiectasis and/or honeycombing, and compared these to Idiopathic Pulmonary Fibrosis (IPF) patients and age-matched controls without respiratory disease. We performed single-cell RNA-sequencing and multiplex immunostaining on peripheral blood mononuclear cells collected at the COVID-19 patients’ first visit after ICU discharge. Six months later, symptoms, restriction and PF improved in some (Early-Resolving COVID PF), but persisted in others (Late-Resolving COVID PF). Results: Circulating monocytes were significantly reduced in Late-Resolving COVID PF patients compared to Early-Resolving COVID PF and non-diseased controls. Monocyte abundance correlated with pulmonary function tests FVC and DLCO. Differential expression analysis revealed MHC-II class molecules were upregulated on the CD8 T cells of Late-Resolving COVID PF patients but downregulated in monocytes. IPF patients had a similar decrease in monocyte abundance and marked decrease in monocyte HLA-DR protein expression compared to Late-Resolving COVID PF patients. Conclusion: Circulating monocyte abundance may distinguish between patients whose post-COVID PF resolves or persists. Furthermore, fibrotic progression coincided with decreases in HLA-DR expression on monocytes, a phenotype previously associated with dampened antigen stimulation and severe respiratory failure.

## Introduction

Over 607 million cases of COVID-19 have been documented worldwide, and up to 30% of patients experience persistent symptoms months after illness^1–4^. Pulmonary fibrosis is common in COVID-19 patients following hospitalization in the intensive care unit (ICU) and mechanical ventilation. 27% of computed tomography (CT)-scanned patients have evidence of fibrosis during hospitalization, which increases to 33% six months after illness^5,6^. The mechanisms that govern the resolution or persistence of pulmonary fibrosis associated with severe COVID-19 are largely unknown. There is a need to define the development of pulmonary fibrosis after infection with SARS-CoV-2 and to determine whether COVID-associated pulmonary fibrosis (COVID PF) is similar to progressive pulmonary fibrosis^7^.

Recent evidence indicates that abnormal immune function plays a significant role in COVID-19 severity^8–11^. For example, SARS-CoV-2 causes a robust inflammatory response within the lung that can lead to the development of acute respiratory distress syndrome (ARDS), tissue damage, and long-term respiratory dysfunction^12–15^. A growing number of studies have begun to characterize COVID PF^16–20^, but further research is critically needed to identify biomarkers that predict poor outcomes within COVID PF and inform treatment plans. Peripheral immune cells are an ideal diagnostic tool as post-COVID-19 sequelae often manifests as a systemic disorder. Furthermore, profiling peripheral blood mononuclear cells (PBMCs) has yielded insights into immune dysregulation and markers that predict disease outcomes in patients with idiopathic pulmonary fibrosis (IPF), the most common fibrotic lung disease^21–25^, setting a precedent that similar methodologies could be applied to COVID PF.

Here, we identify a cohort of COVID-19 patients who had restrictive lung physiology and early CT scan changes consistent with fibrosis more than one month after acute SARS-CoV-2 infection symptoms had resolved. At six-month outpatient follow-up visit, the cohort diverges into two groups: Patients whose physiologic restriction and early fibrotic changes resolved, which we termed “Early-Resolvers” (ER COVID-PF), and those with persistent restriction and pulmonary fibrosis, whom we termed “Late Resolvers” (LR COVID-PF) (Figure 1A). The objective of our study was to first define immune features that discriminated LR COVID-PF from ER COVID-PF patients by performing single-cell RNA Sequencing and multiplex immunostaining analysis of PBMCs, and to second compare the resultant cellular and molecular signatures with IPF.

**Figure 1.**
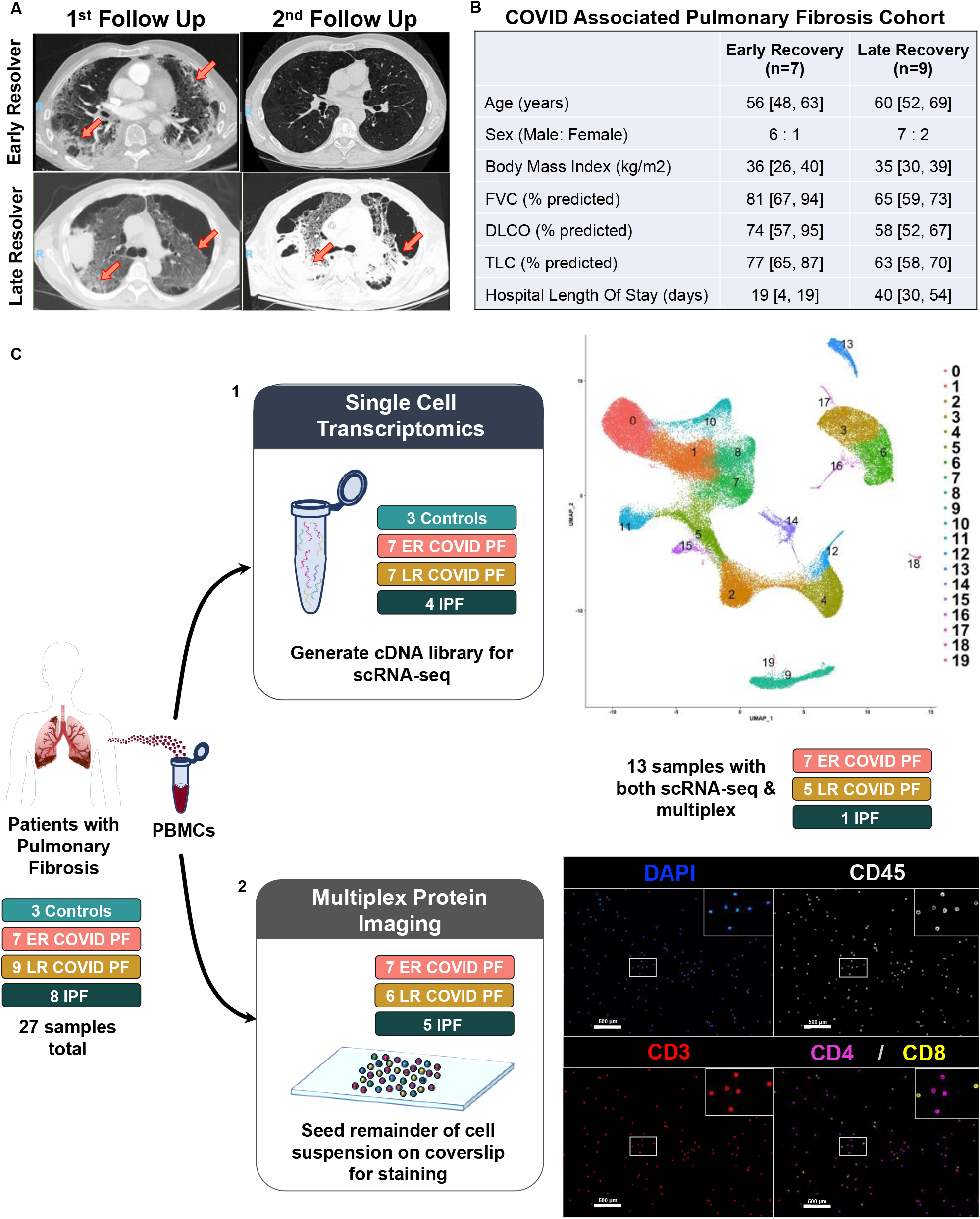
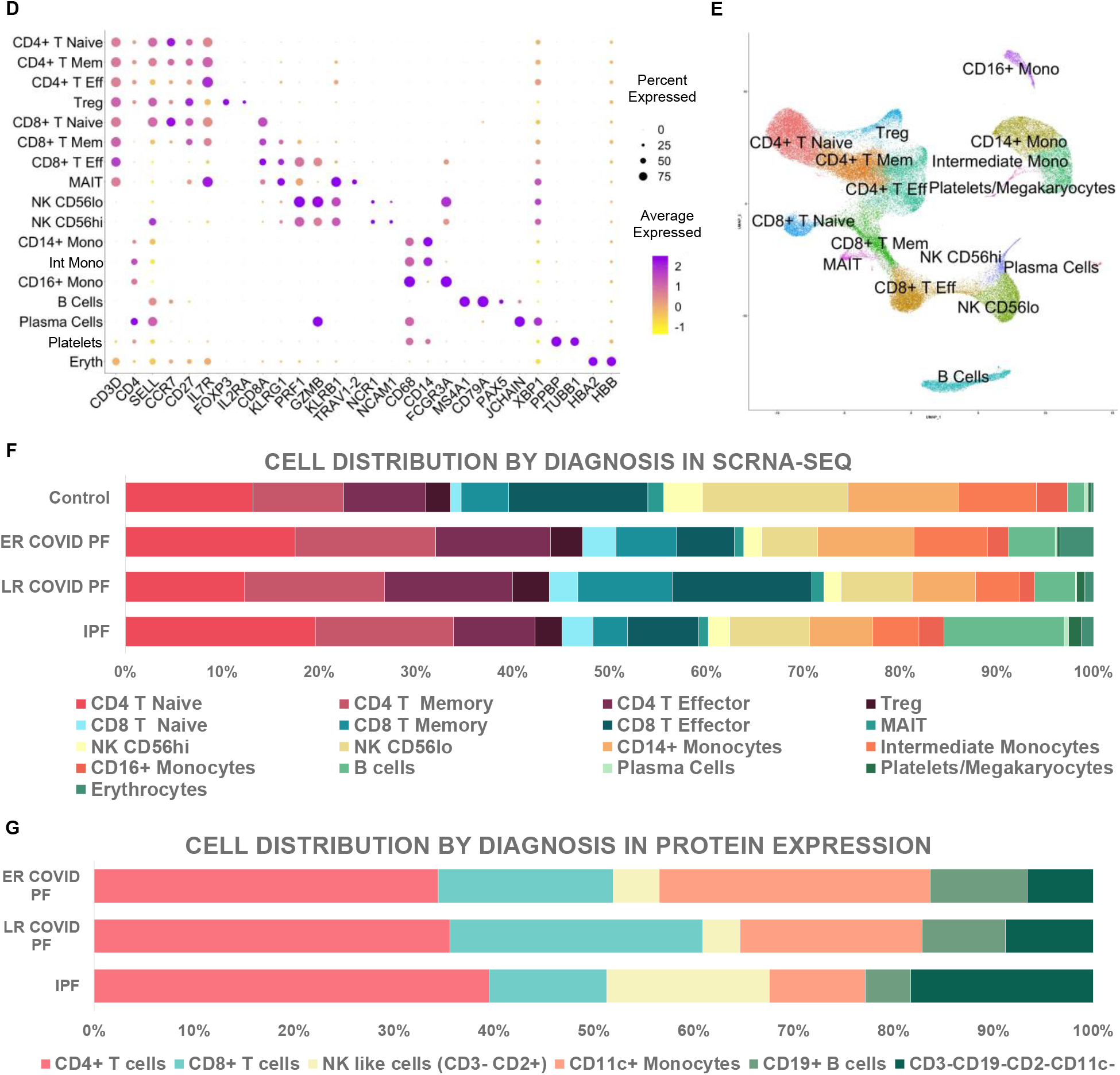
Clinical features of patients with COVID associated pulmonary fibrosis and study design. *(A)* CT images of COVID survivor with resolving pulmonary fibrosis (top row) and with persistent fibrosis (bottom row). Left images represent abnormal CT findings observed when first admitted to the post-COVID clinic more than one month post-infection. Right images denote either the resolution or persistence of abnormal findings 6+ months after infection. Abnormal findings such as grey opacities and reticulation are indicated by orange arrows. *(B)* Table detailing clinical characteristics of early and late resolvers within COVID associated pulmonary fibrosis patients. *(C)*. Schematic of multi-omic study design where *(1)* depicts scRNA-seq processing and preliminary UMAP, and *(2)* depicts multiplex imaging workflow and representative image of stained PBMCs using PhenoCycler to identify T cells. *(D)* Dot plot denoting expression of marker genes used to identify 16 PBMC subpopulations as well as plateletes and erythrocytes. (*E)* UMAP of cell clusters from integrated data of control, COVID, and IPF PBMCs generated through scRNA-seq in Seurat (21 samples total). *(F)* Stacked bar chart depicting relative cell abundances within each group (Control, ER COVID PF, LR COVID PF, and IPF) for the 16 subclusters identified by scRNA-seq. *(G)* Stacked bar chart depicting relative cell abundances within each treatment (ER COVID PF, LR COVID PF, and IPF) for the 6 subpopulations identified by immunostaining and imaging with the PhenoCycler. *\*PBMC = peripheral blood mononuclear cell; scRNA-seq = single cell RNA sequencing; ER COVID PF = “Early-Resolvers”; LR COVID PF = “Late Resolvers”; IPF = Idiopathic Pulmonary Fibrosis; NK = Natural Killer Cells; FVC = Force Vital Capacity; DLCO = Diffusing Capacity for Carbon Monoxide; TLC = Total Lung Capacity; Eff = Effector; Mem = memory; Reg = regulatory*.

## Methods

Single-cell RNA sequencing (scRNAseq) was performed on cryopreserved PBMCs from four IPF and 14 COVID-19 associated pulmonary fibrosis (COVID PF) patients at the University of Virginia. Patients with IPF donated PBMCs in the outpatient setting while in a stable clinical state. Three control PBMC samples from age-matched patients with no known pulmonary disease were prepared and sequenced at the Mayo Clinic (IRB #:19-012187). We recruited a subset of patients from the University of Virginia COVID-19 survivor clinic with restrictive lung physiology on pulmonary function tests (PFTs) and features consistent with pulmonary fibrosis on chest CT^26^. Radiographic features indicative of possible development pulmonary fibrosis included bilateral reticulation, traction bronchiectasis, and/or honeycomb change in peripheral and basilar distribution, similar to the presently recognized progressive pulmonary fibrosis clinical radiologic phenotype, and similar to previously defined COVID-19 pulmonary fibrosis characteristics^7^. PFT and chest imaging was performed in association with the patient’s first visit to the outpatient COVID-19 survivor clinic. Early- and Late-Resolvers were identified by comparing chest imaging and PFT values from the patient’s first visit to their subsequent testing. Patients with COVID-19 associated pulmonary fibrosis (COVID PF) were followed for 6 months or until the patient improved. COVID PF patients were age-matched to IPF patients to control for age-related differences in peripheral immune signatures.

### Single cell RNA sequencing and data preparation

Peripheral blood samples were prepared using the 10x Genomics Fresh Frozen Human PBMC protocol and submitted to the UVA Genomics Core for library preparation.

ScRNA-seq data were filtered for dead cells, doublets, and red blood cells by excluding cells with greater than 5% mitochondrial genes and less than 500, but no more than 2500 genes. Samples underwent normalization, integration, dimensional reduction, and further downstream analysis using the standard Seurat workflow with the Seurat v.4.0.3 package. Further details regarding parameters for clustering, the differential expression analysis, and statistics can be found in the online supplement.

### Multiplex immunostaining

Protein expression was measured using multiplex imaging which can utilize samples with low cell count. PBMCs from the same suspension processed for scRNA-seq were seeded on a poly-L-lysine coated coverslip and prepared for multiplex imaging on the PhenoCycler (Akoya Biosciences) according to the manufacturer’s protocol. Six ER COVID-PF, seven LR COVID-PF, and five IPF samples were immunostained and quantified. Of those, six ER COVID PF, five LR COVID PF, and one IPF samples were also processed for sc-RNA-seq.

## Results

### COVID Associated Pulmonary Fibrosis cohort clinical features

16 COVID-19 survivors were recruited at their first outpatient follow up after recovery. All sought care for persistent symptoms of dyspnea and fatigue, and none had known pulmonary fibrosis prior to COVID-19. All but 2 patients had required hospitalization for COVID-19. Although most of our cohort were ICU survivors, two patients were evaluated in the COVID-19 survivor clinic with persistent impaired respiratory function despite never requiring hospitalization. This supports a growing body of evidence that patients with mild COVID cases also have the potential to develop long term symptoms^27,28^. Nevertheless, both non-hospitalized patients showed an early recovery trajectory, maintaining that patients with increased age and prolonged hospitalization are most at risk^3^. 10 patients received mechanical ventilation, 1 received non-invasive positive pressure ventilation, and 3 received high-flow nasal cannula oxygen. Mean time on mechanical ventilation was 30 ± 18 days, defined as either time of intubation to successful extubation or the ability to perform tracheostomy collar without positive pressure ventilation 24 hours per day when tracheostomy with prolonged ventilator weaning was required. Prospective studies have indicated that non-invasive and invasive mechanical ventilation are independent risk factors of fibrotic lung changes following COVID-19^3,18^. Supporting this notion, eight out of nine (88.8%) LR COVID PF patients required mechanical ventilation as opposed to two out of seven (28.5%) ER COVID PF.

Hospital length of stay was prolonged (Figure 1B), with many patients requiring discharge to skilled nursing facilities for rehabilitation. Mean time since testing positive for COVID-19 was 6 ± 3.5 months. Three patients still required supplemental oxygen at levels between 2 and 6 liters via nasal cannula. All patients were followed prospectively for six months or more. There was no significant difference in age, sex, or body mass index between patients who developed early versus late recovery trajectories (Figure 1B), and no significant difference in medical comorbidities (Table E1). ER COVID PF patients showed improvement in their PFT and chest imaging, with 6 of the 7 patients having normal range PFT at their last follow up, at mean 9.5 months after testing positive for COVID-19. One ER COVID PF patient had persistent mild restriction on PFT, but no imaging findings to suggest interstitial fibrosis. Nine patients, all of whom required ICU level hospitalization, showed a late recovery trajectory. At mean follow up 11 months after testing positive for COVID-19 and 7 months after hospital discharge, 6 of these 9 patients had persistent restrictive physiology. All had chest imaging showing bilateral fibrotic changes.

### Idiopathic Pulmonary Fibrosis and Control cohort clinical features

8 IPF patients comparable in age were also selected as a positive control of chronic progressive pulmonary fibrosis. Mean age was 65, mean BMI was 34, and 7 of the 8 patients were men, matching the demographics and body mass of the COVID-19 cohort (Table E1). PFTs measured at their clinically stable outpatient visit showed mean FVC 64% of predicted and DLCO 46% of predicted. Six of 8 IPF patients used supplemental oxygen, ranging between 2 and 8 liters. In addition, 3 age-matched patients without known pulmonary disease were sequenced as a non-pulmonary disease control (Mean age = 69, All male).

### Single-cell RNA sequencing

To profile the peripheral immune response in post-COVID pulmonary fibrosis, we generated two rich and complementary datasets using PBMC samples (Figure 1C). For in-depth characterization of transcriptional differences, we performed scRNA-seq on PBMCs from 21 subjects (seven ER COVID-PF, seven LR COVID-PF, four IPF, and three age-matched non-diseased controls). The data were integrated to yield a combined 71,574 cells. Nineteen cell clusters were identified after dimensionality reduction by uniform manifold approximation and projection (UMAP). Erythrocyte clusters were removed, and two CD4+ T effector cell clusters were merged, yielding 16 distinct immune cell subpopulations (69,868 cells), which were identified and manually annotated using established markers (Figure 1D-E)

### Multiplex immunostaining

To complement this sequencing dataset with protein-level data, we generated a 13-antibody panel (Figure 1C2, Figure E1, Table E2) and immunostained PBMC samples from 18 subjects (six ER COVID PF, seven LR COVID PF, and five IPF). After blinded gating, B cell, Monocyte, NK-like, CD4+ and CD8+ T cell subpopulations were identified (Figure E1). Eleven COVID samples and one IPF sample underwent both scRNA-seq and multiplex immunostaining (Table E3).

All major subpopulations identified by immunostaining and all 16 subpopulations identified by scRNA-seq were present in each patient sample, indicating data consistency (Figure 1F-G). These datasets enabled comparisons of immune cell abundance and gene expression among patients with ER and LR COVID PF, as well as patients with IPF and age-matched controls.

### Relative cell abundances vary between Early- and Late-Resolving COVID PF

First, we examined whether the relative abundances of circulating immune cells differ in ER and LR COVID PF (Figure 2A-I). Compared to controls, both ER and LR COVID PF patients had significantly fewer plasma cells (p = 0.0378 and p = 0.0044, respectively) and NK-like cells (p = 0.0227, p = 0.0473, Figure 2I, E). There were no differences in CD4+ T cell abundance between groups, however we saw a unique decrease in the ratio of naïve to effector (where effector is the sum of regulatory, memory, and effector subpopulations) within the LR COVID PF group (p=0.0601). LR COVID PF patients had increased CD8+ T cells compared to ER COVID PF patients, showing a 1.6-fold increase in CD8+ T memory cells. There were trending increases in effector and total CD8+ T cells within the LR COVID PF group at the transcript and protein level that did not reach significance, and there was no significant difference in the ratio of naïve to effector CD8+ T cells (sum of memory and effector) (Figure 2B, D). Most notably, LR COVID PF patients showed significant decreases in all monocyte populations compared to both ER COVID PF patients and controls (Figure 2G-H). For rigor, we tested whether time from initial COVID positivity to sample collection was correlated with monocyte abundance and found no correlation. Quantification of cellular abundance by multiplex immunostaining corroborated that the relative abundance of CD11c+ monocytes to all PBMCs was significantly lower in patients with LR COVID PF compared to ER COVID PF (p = 0.0036, Figure 2H).

**Figure 2.**
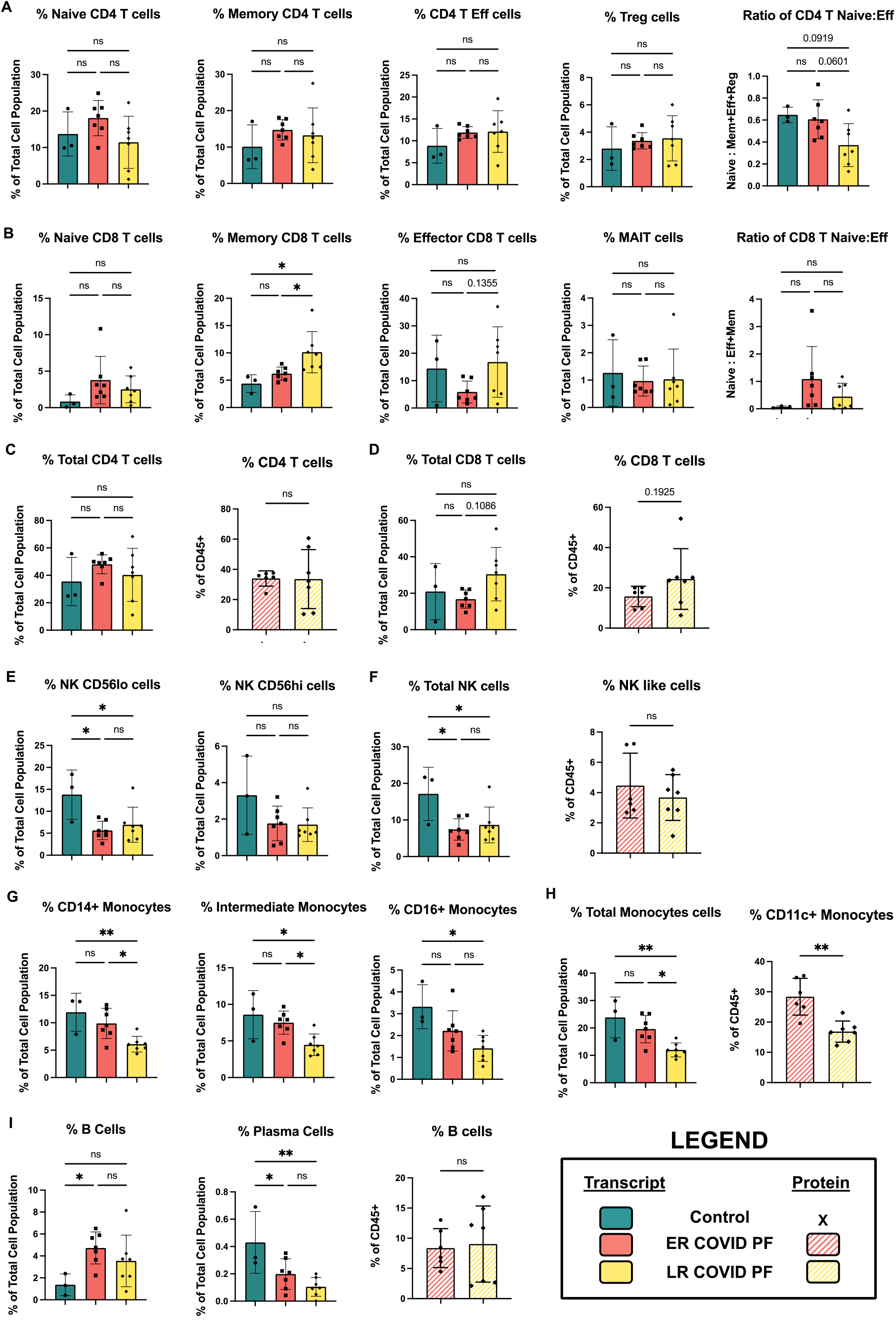
Cell abundances vary between ER and LR COVID PF. *(A)* ScRNA-seq relative abundances of CD4+ T cell populations and ratio of naïve to effector (defined as the sum of effector, memory, and regulatory cells here) in Control, ER COVID PF, and LR COVID PF. *(B)* ScRNA-seq relative abundances of CD8+ T cell populations and ratio of naïve to effector (defined as the sum of effector, memory, and regulatory cells here) in Control, ER COVID PF, and LR COVID PF. Relative total abundances of CD4+ T cells *(C)* and CD8+ T cells *(D)* identified by scRNA-seq, left (solid), and protein on PhenoCycler, right (striped). *(E)* ScRNA-seq relative abundances of natural killer subpopulations. *(F)* Relative abundance of total natural killer cell population identified by scRNA-seq, left (solid), and protein on PhenoCycler, right (striped). *(G)* ScRNA-seq relative abundances of monocyte subpopulations. *(H)* Relative abundance of total monocyte cell identified by scRNA-seq, left (solid), and protein on PhenoCycler, right (striped). *(I)* ScRNA-seq relative abundances of B and plasma cell subpopulations (solid) and protien quantification of total CD19+ B cell population on PhenoCycler, right (striped).ScRNA-seq data in *A-I* was tested for significance using an ordinary one-way ANOVA (parametric, equal SD) with Tukey’s multiple comparison test where the mean of each group (ER COVID PF, LR COVID PF, and IPF) was compared to eachother. Significance for protein quantification in *C-I* was tested using Welch’s t-test (parametric, unpaired) to compare ER COVID PF and LR COVID PF.

### Depletion of circulating monocytes in COVID PF patients correlates with decreases in pulmonary function

The decrease in relative monocyte abundance within the LR COVID PF group was the only difference found to be significant at both the transcript and protein level. We therefore sought to determine whether monocyte abundance was correlated with pulmonary function in post-COVID PF patients. The relative abundances of CD14+ monocytes, intermediate monocytes (CD14+ CD16+), and total monocytes were each positively correlated with both forced vital capacity (% predicted FVC, R^2^ = 0.52, 0.60, 0.55, respectively) and the diffusing capacity for carbon monoxide (% predicted DLCO, R^2^ = 0.50, 0.53, 0.51, respectively) (Figure 3A-B).

**Figure 3.**
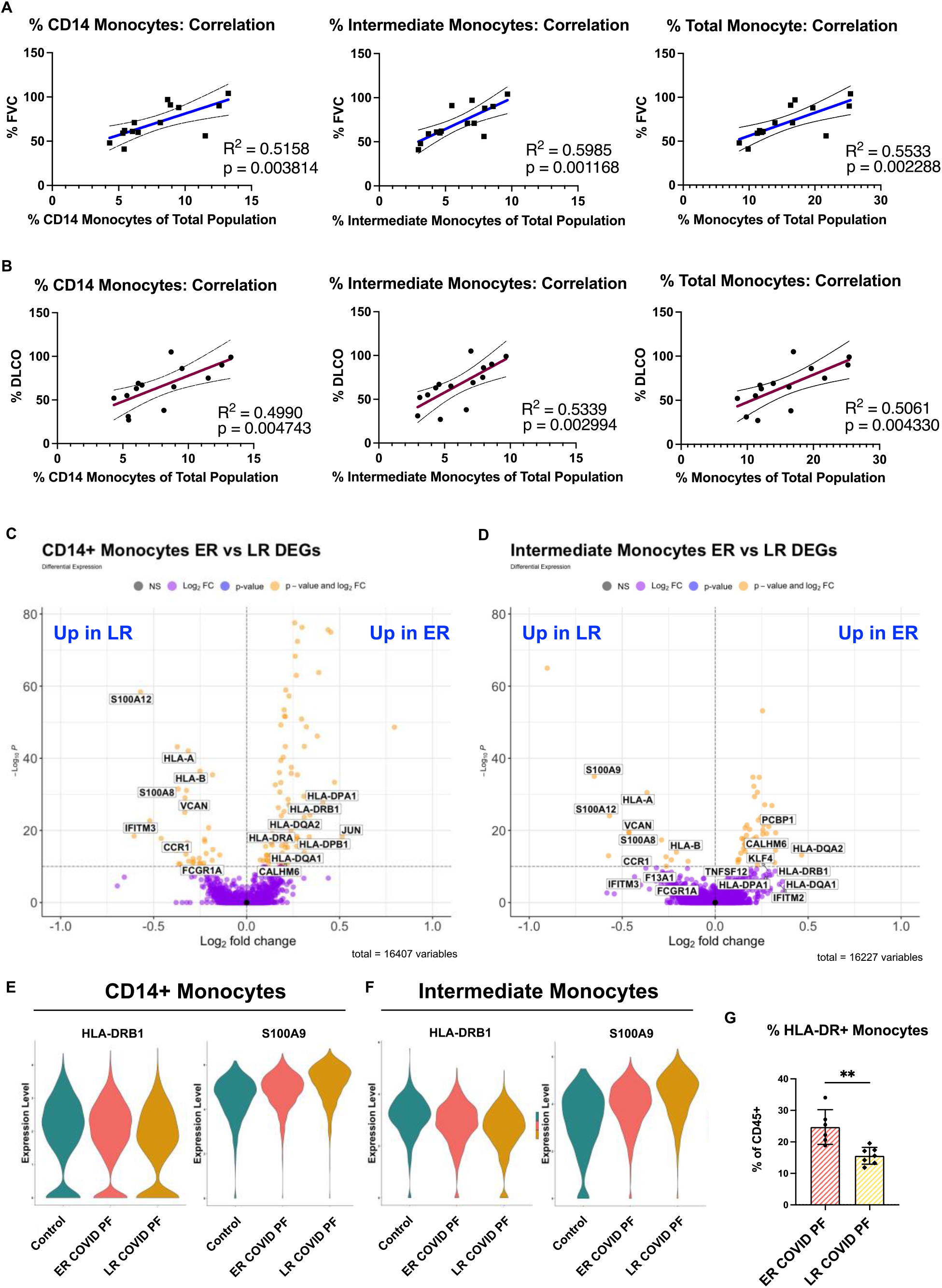
Decreased monocyte abundance correlates with decreased lung function in COVID associated pulmonary fibrosis. Pearson correlation of % Force Vital Capacity (FVC) *(A)* and % Diffusing Capacity for Carbon Monoxide (DLCO) *(B)* compared to relative abundance of CD14+, intermediate, and total mkonocytes from scRNA-seq. Volcano plot showing differentially expressed genes between ER and LR COVID PF generated from MAST analysis for CD14+ monocytes *(C)* and intermediate monocytes *(D)* where positive log2FC values represent genes upregulated in ER COVID PF relative to LR COVID PF and negative log2FC represent genes upregulated in LR COVID PF relative to ER COVID PF. *(E-F)* Violin plots showing gene expression of MHC-II molecule, HLA-DRB1, and the alarmin, S100A9, between control, ER COVID PF, and LR COVID PF. *(G)* Quantification of the percent of monocytes expressing HLA-DR at the protein level from imaging with the PhenoCycler. Significance for protein quantification in *G* was tested using Welch’s t-test (parametric, unpaired).

To evaluate whether there were differences in the transcriptome of ER and LR COVID PF monocytes, differential expression analysis was performed. Alarmins, such as S100A12, S100A9, and S100A8, were among the most significantly enriched genes within the monocytes of patients with persistent fibrosis (Figure 3E-F). Differential expression analysis also revealed downregulation of MHC class II molecules, such as HLA-DPA1, HLA-DPB1, HLA-DRB1, and HLA-DRA, in circulating CD14+ and intermediate monocytes from LR COVID PF in comparison to both ER COVID PF and non-diseased controls (Figure 3C-D). Consistent with these transcriptomic results, quantification of HLA-DR protein revealed a significant 37% decrease (p = 0.0075) in the proportion of MHC-II molecule expressing circulating monocytes in patients who fail to resolve their pulmonary fibrosis compared to ER COVID PF (Figure 3G). Our data are consistent with previous studies that have shown increased alarmins and decreased MHC-II molecule expression on monocytes in severe COVID verses mild disease^12,29–33^, while also establishing that these markers can be used to further differentiate patient outlook within severe COVID, specifically pulmonary fibrosis associated with COVID.

### Late-Resolving COVID Associated Pulmonary Fibrosis patients show pro-longed CD8+ T cell activation and increases in CD4+ T effector populations

To investigate whether T cell-associated mechanisms differentiated ER and LR COVID PF, we identified unique transcript and protein signatures among lymphocytes. CD8+ T effector cells had the highest number of differentially expressed genes of all PBMC subpopulations, suggesting elevated activation status (148 DEGs with p_adj_-value < 0.10 and 77 DEGs after adjusting for time between initial infection and sample collection) (Tables E4). Contrary to LR COVID PF monocytes, HLA-DRA, HLA-DPA1, and HLA-DRB5 were among the top DEGs upregulated in LR COVID PF CD8+ T effector cells when compared to ER COVID PF and non-diseased control (p_adj_-value < 1.0E-10) (Figure 4A-B). These increases persisted when controlling for time from initial COVID positivity to sample collection (Tables E4). Immunofluorescent staining confirmed that CD8+ T cells of LR COVID PF patients had nearly a 3-fold increase in HLA-DR protein expression as well 4-fold increase in the proportion of CD8+ T cells that co-express HLA-DR+ and CD38+, a phenotype associated with lymphocyte activation (Figure 4C). Given that PBMCs were collected months after initial infection, these results suggest LR COVID patients exhibit prolonged immune activation, reminiscent of chronic inflammation.

**Figure 4.**
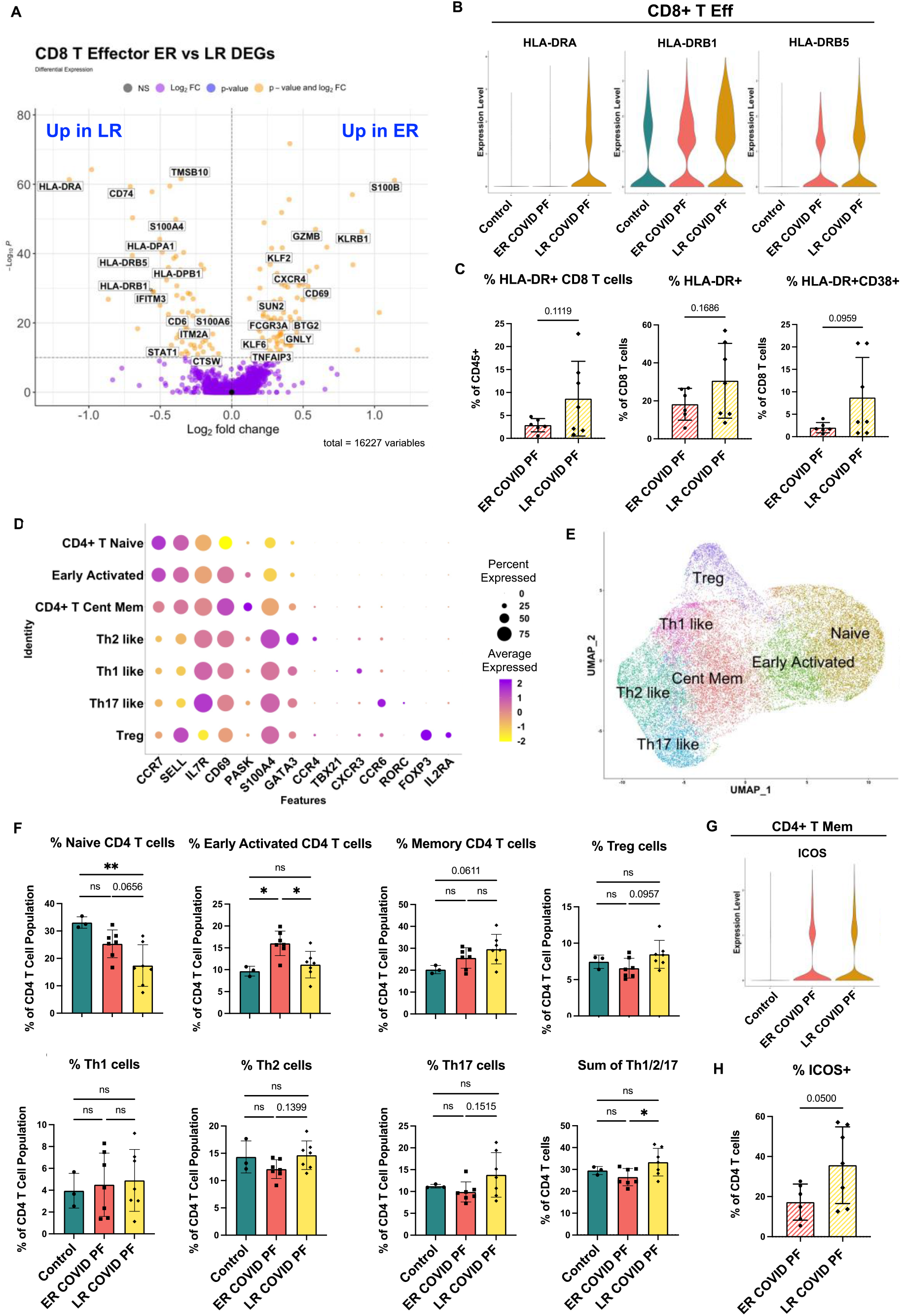
Late-Resolving COVID associated pulmonary fibrosis exhibits molecular hallmarks of prolonged T cell activation. *(A)* Volcano plot showing differentially expressed genes between ER and LR COVID PF generated from MAST analysis for CD8+ T effector cells where positive log2FC values represent genes upregulated in ER COVID PF relative to LR COVID PF and negative log2FC represent genes upregulated in LR COVID PF relative to ER COVID PF. *(B)* Violin plots showing gene expression of MHC-II molecules in CD8+ T effector cells between control, ER COVID PF, and LR COVID PF. *(C)* Quantification of the percent of CD8+ T cells in ER and LR COVID PF expressing HLA-DR as a percent of the total population as well as HLA-DR and CD38 as a percent of T cells at the protein level from imaging with the PhenoCycler. *(D)* Dot plot denoting expression of marker genes used to identify CD4+ T cell subpopulations. *(E)* UMAP of subclusters generated from re-clustering CD4+ T cells. *(F)* ScRNA-seq relative abundances of CD4+ T relative to the total CD4+ T cell population for control, ER COVID PF, and LR COVID PF. *(G)* Violin plot showing gene expression of ICOS in CD4+ T memory cells between control, ER COVID PF, and LR COVID PF. *(H)* Quantification of the percent of CD4+ T cells in ER and LR COVID PF expressing ICOS at the protein level from imaging with the PhenoCycler. Significance for protein quantification in *C&H* was tested using Welch’s t-test (parametric, unpaired). *F* significance was tested using an ordinary one-way ANOVA (parametric, equal SD) with Tukey’s multiple comparison test where the mean of each group was compared to eachother.

To execute an unbiased analysis of the changes between ER and LR COVID PF gene expression within CD4+ and CD8+ T effector cells, we implemented the Model-based Analysis of Single-cell Transcriptomics (MAST) test and clusterProfiler package to perform gene set enrichment analysis (GSEA)^34–36^. In alignment with our hypothesis that LR COVID PF patients exhibit prolonged T cell activation, eight T cell receptor signaling pathways were enriched in LR COVID PF patients compared to controls (Tables E7, Figure E3). Furthermore, CD8+ T effector cells in both ER and LR COVID PF were positively enriched for gene sets involving Proinflammatory and Profibrotic Mediators, Cytokine-cytokine Receptor Interactions, and the Network Map of SARS-CoV2 Signaling Pathway when compared to control. Within ER and LR COVID PF, we also observed enrichment of multiple IL-1 and CD40 pathways as well as the Senescence Associated Secretory Phenotype (SASP). Notably, Allograft Rejection and Interferon Gamma (IFN-γ) Signaling were significantly enriched in LR COVID PF when compared to control and ER COVID PF. This corroborates other studies which have found increased IFN-γ in severe COVID^29,37,38^. Both CD8+ and CD4+ T effector cells in ER and LR COVID PF were positively enriched for IL-10, IL-18, and multiple MAPK and Toll Like Receptor Signaling pathways compared to control (Table E7). Interestingly, ER and LR COVID PF patients were negatively enriched for Extracellular Matrix Organization in CD8+ T effector cells and ECM Regulators in CD4+ T effector cells in comparison to control, suggesting that T cell mediated ECM regulation may be decreased in COVID PF. We observed upregulation of the TGF-β pathway, a key driver of fibrosis, in ER and LR COVID PF CD4+ T effector cells (Table E7).

To further identify classical T helper cell subtypes, we reclustered all CD4+ T cells, yielding seven distinct populations: Naïve, Early Activation (denoted by high expression of naïve markers with moderate expression of activation markers), Central Memory, and several distinct effector populations (Th1, Th2, Th17, and Treg) that expressed canonical markers (Figure 4D). Corroborating results in Figure 1F, LR COVID PF patients had significantly fewer naïve CD4+ T cells than both control and ER COVID PF (Figure 4F), and had a smaller Early Activated population compared to ER COVID PF. In contrast, central memory, Treg, and effector (sum of Th1, Th2, and Th17) CD4+ T cells were increased in LR COVID PF. These results suggest that the CD4+ T helper response in LR COVID PF has been skewed to an active effector/memory phenotype. In support of this notion, we found a significant increase in the protein expression of the co-stimulatory molecule ICOS (Figure 4H), as well as increased protein expression of PD1 and CD69 on CD4+ T cells of LR COVID PF patients compared to those of ER COVID PF patients (Figure E2). To determine which CD4+ T cell population was expressing ICOS, we queried our scRNA-seq data and detected ICOS on the CD4+ T memory population (Figure 4G). As ICOS is rapidly expressed after T cell receptor engagement and broadly expressed in activated T cells, this data aligns with our GSEA and CD4+ T cell abundance findings which suggested a more activated phenotype within LR COVID PF patients.

### LR COVID PF patients maintain a perpetual T cell activation response in comparison to both their ER COVID PF counterparts and IPF

To evaluate whether the immune response in LR COVID PF has similarities to other chronic interstitial lung diseases, we next compared the immune signatures of LR COVID PF to IPF. Unlike LR COVID PF, there was no change in the total number of T cells or ratio of naïve to effector T cells in IPF when compared to control (Figure 5A-D). Further investigation using our re-clustered CD4+ T cell dataset showed IPF patients had more naïve cells and a significant decrease in all effector cell populations, except Th1-like cells, compared to LR COVID PF (Figure 5I).

**Figure 5.**
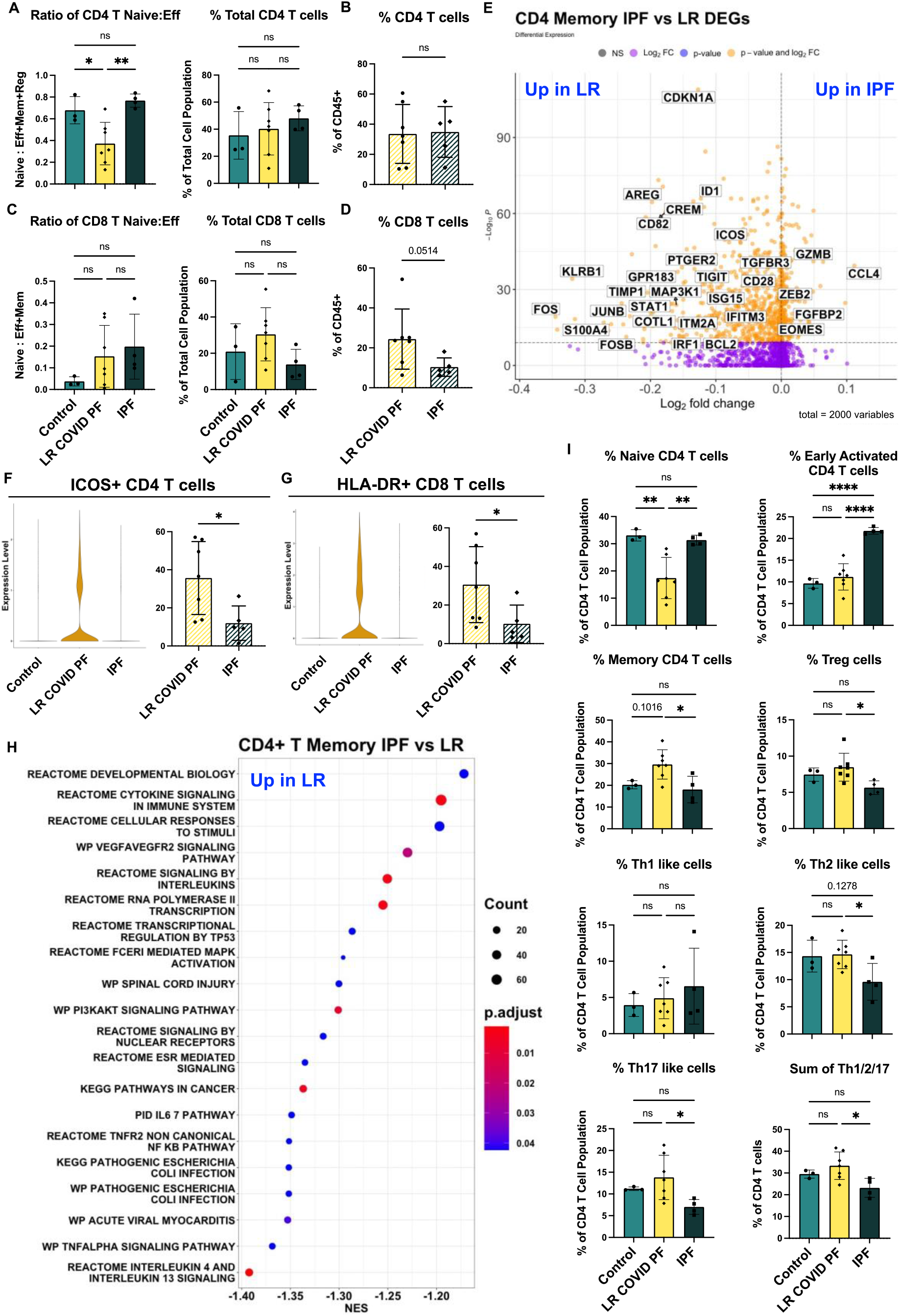
Adaptive immune cells differentiate IPF and COVID PF. *(A)* ScRNA-seq data of CD4+ T cells showing the ratio of naïve to effector (defined as the sum of effector, memory, and regulatory cells here) and relative total abundance of CD4+ T cells in Control, LR COVID PF, and IPF. *(B)* Protein qunitifcation of relative total abundance of CD4+ T cells in LR COVID PF and IPF. *(C)* ScRNA-seq data of CD8+ T cells showing the ratio of naïve to effector (defined as the sum of effector and memory cells here) and relative total abundance of CD8+ T cells in Control, LR COVID PF, and IPF. *(D)* Protein qunitifcation of relative total abundance of CD8+ T cells in LR COVID PF and IPF.*(E)* Volcano plot showing differentially expressed genes between IPF and LR COVID PF generated from MAST analysis for CD4+ T memory cells where positive log2FC values represent genes upregulated in IPF relative to LR COVID PF and negative log2FC represent genes upregulated in LR COVID PF relative to IPF. *(F)* Violin plots showing gene expression of ICOS in CD4+ T memory cells between control, ER COVID PF, and LR COVID PF and quantification of the percent of CD4+ T cells expressing ICOS at the protein level from imaging with the PhenoCycler. *(G)* Violin plots showing gene expression of MHC-II molecules (HLA-DR) in CD8+ T effector cells between control, ER COVID PF, and LR COVID PF and quantification of the percent of CD8+ T cells expressing HLA-DR at the protein level from imaging with the PhenoCycler. *(H)* Dot plot depicting GSEA analysis results of the top 20 pathways enriched in LR COVID PF compared to IPF, where negative NES are enriched in LR COVID PF relative IPF. *(I)* ScRNA-seq relative abundances of CD4+ T relative to the total CD4+ T cell population for control, LR COVID PF, and IPF. *A, C, & I* significance was tested using an ordinary one-way ANOVA (parametric, equal SD) with Tukey’s multiple comparison test where the mean of each group was compared to eachother. Significance for protein quantification in *B, D, F, and G* was tested using Welch’s t-test (parametric, unpaired).

To determine which genes were most significantly up or down-regulated in LR COVID PF compared to IPF, we performed DEG analysis on all 16 scRNA-seq clusters. We found CD4+ T memory cells had the most DEGs (771) where p<0.10. Notably, genes involved in interferon signaling (e.g., ISG15, IRF1, IFITM3), which has been highlighted as a key pathway in severe COVID^29,37,38^, were upregulated in CD4+ T memory cells of LR COVID PF patients (Figure 5E). In accordance with previous IPF literature^39–41^, we find the CD4+ T memory cells in IPF patients were characterized by gene expression patterns associated with senescence and exhaustion, such as ZEB2, EOMES, and decreased CD28 expression^42–44^. Previously, loss of CD28 and ICOS correlated with reduced transplant-free survival in IPF^39^. Here, we observed reductions in both CD28 and ICOS in IPF compared to LR COVID PF at the transcript level (Figure 5E). Expression of ICOS protein was also significantly greater (p=0.0188) in CD4+ T cells of LR COVID PF compared to IPF, suggesting that T cell activation is enhanced in LR COVID PF in comparison to control, ER COVID PF, and IPF (Figure 5F). Furthermore, LR COVID PF have increased expression of HLA-DR in CD8+ T effector cells as well as a higher percentage of CD8+ T cells co-expressing HLA-DR+ and CD38+(Figure 5G). GSEA of CD4+ T memory cells in IPF versus LR COVID PF further demonstrated that LR COVID PF patients exhibit signs of potentiated inflammation as indicated by enrichment of TNFα, cytokine, and multiple interleukin signaling pathways (Figure 5H). These data suggest T cells from LR COVID PF patients are more active than their IPF counterparts, which express more senescence and exhaustion markers.

### Monocyte abundance and HLA-DR expression link Late-Resolving COVID Associated Pulmonary Fibrosis and IPF

Despite differences in the adaptive immune response, we observed similarities in the innate immune responses of patients with LR COVID PF and IPF (Figure 6A-F); scRNA-seq results showed that both conditions exhibited a nearly 50% reduction in monocytes compared to controls (Figure 6E). Quantification by immunofluorescent staining corroborated that LR COVID PF and IPF patients had similarly low monocyte abundances compared to ER COVID PF (p=0.0743, p=0.0138 respectively) (Figure 6F). Strikingly, monocytes from IPF patients expressed significantly lower levels of HLA-DR protein than LR-COVID PF patients when compared to ER COVID PF (p=0.0009, p=0.0265) (Figure 6H). Cross-referencing our scRNA-seq dataset revealed that HLA-DR expression was decreased only on non-classical CD16+ monocytes in IPF, whereas in LR COVID PF the decrease in HLA-DR was observed in the classical CD14+ and intermediate monocytes but not on non-classical CD16+ subpopulation (Figure 6G, Tables E6). These results suggest that reductions in monocyte HLA-DR are a common feature of severe fibrotic lung disease.

**Figure 6.**
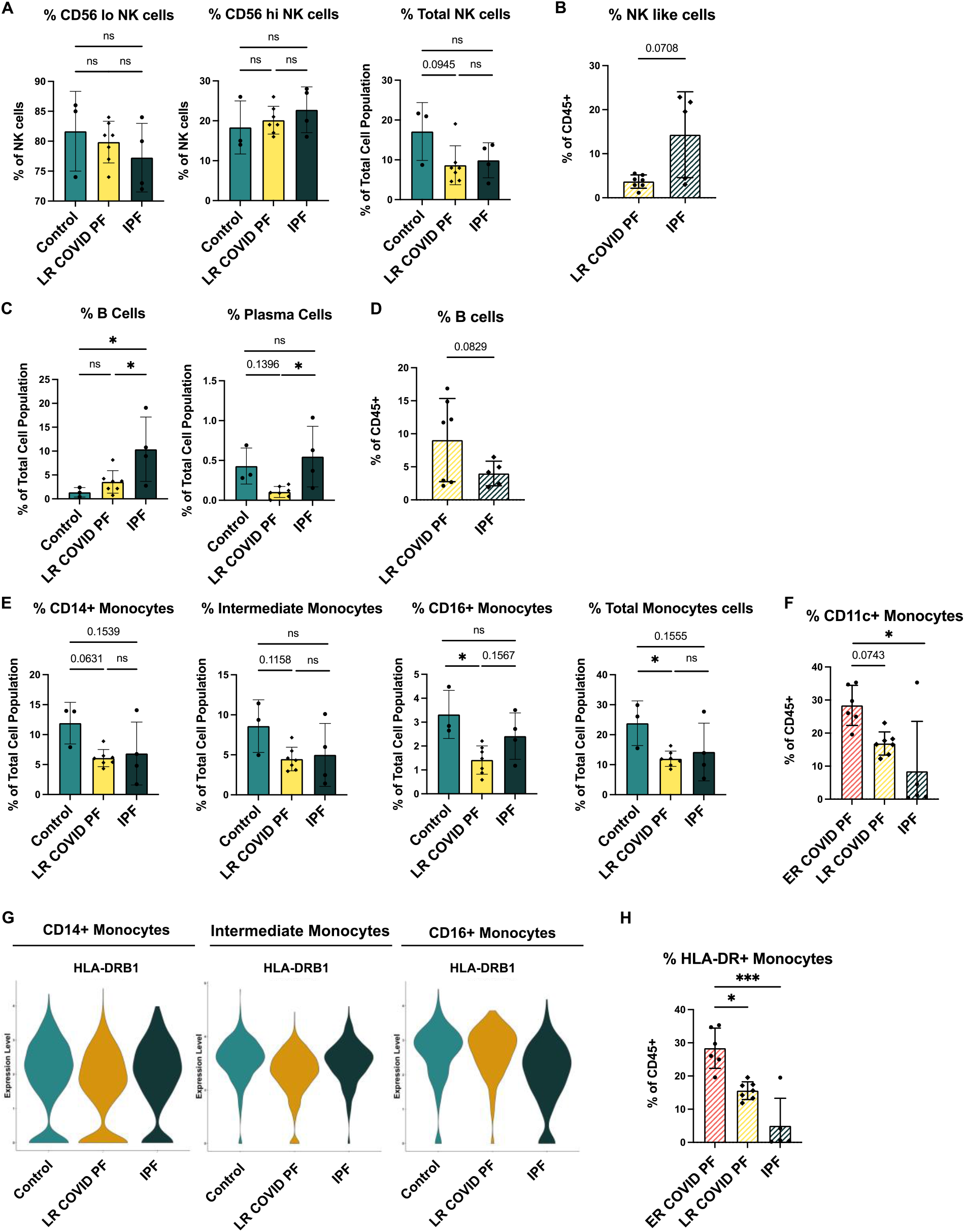
Innate immune cells in LR COVID PF resemble IPF. *(A)* Quantification of scRNA-seq relative abundances of subpopulations and total population of natural killer cells *(A)*, B cells *(C)*, and monocytes *(E)* for control, LR COVID PF, and IPF. *A, C, & E* significance was tested using an ordinary one-way ANOVA (parametric, equal SD) with Tukey’s multiple comparison test where the mean of each group was compared to eachother. Quantification of all natural killer cells *(B) and* B cells *(D)* identified by protein imaging on PhenoCycler. *B&D* significance was tested using Welch’s t-test (parametric, unpaired). Quantification of all monocytes *(F)* and percent of the percent of HLA-DR+ monocytes relative to all CD45+ cells in *(H)* in ER COVID PF, LR COVID PF, and IPF from imaging with PhenoCycler. *F&H* significance was tested using Kruskal-Wallis (nonparametric ANOVA) test with Dunn’s multiple comparison test where the rank mean of LR COVID PF and IPF to the mean of ER COVID PF as control. *(G)* Violin plots showing gene expression of MHC-II molecule between IPF, ER and LR COVID PF. Significance for differential expression gene analysis in *G* was determined by non-parametric Wilcoxon rank sum test.

## Discussion

We examined a unique population of COVID-19 convalescent patients with persistent dyspnea and fatigue, who had abnormal PFTs and imaging suggestive of early pulmonary fibrosis. While some patients clinically improved in the outpatient setting in a matter of months (ER COVID PF), others did not (LR COVID PF). Here, we used a multi-omic approach to analyze blood samples collected more than one-month post-infection but before these two cohorts clinically diverged, and we uncovered that immune cell composition and gene expression significantly differed between ER and LR COVID PF patients.

A key finding of this study is LR COVID PF patients had significantly fewer monocytes than ER COVID PF patients and controls. Our study is the first to identify that decreased monocyte abundance correlates with impaired pulmonary function in COVID-associated pulmonary fibrosis. These findings are consistent with reports that severe COVID patients exhibit a decrease in monocytes^30,31,45^, but demonstrate monocyte depletion is a distinguishing characteristic within severe COVID. We also find that monocytes of LR COVID PF patients expressed lower levels of MHC class II molecules. CD16+ monocytes are more mature and express more HLA-DR in comparison to CD14+ monocytes, therefore reductions in HLA-DR+ CD14+ monocytes has been associated with mobilization of immature monocytes from the bone marrow for emergency myelopoiesis^46^ which we observe here and others reported as a marker of severe COVID-19^45,47^. Loss of HLA-DR on monocytes is also an established marker of immunosuppression, so these findings may also suggest dampening of antigen-mediated stimulation and inhibition of antigen-specific T cell responses as has been shown in sepsis^48–50^. Aligned with this hypothesis, Arunachalam et al. reported functional suppression of COVID-19 monocytes compared to healthy controls^33^. Decreases in the MHC class II molecule HLA-DR on monocytes was also associated with severe respiratory failure in COVID-19 pneumonia^12^, immunosuppression^51^, and decreased oxygen saturation in severe COVID-19^52^. Of note, the cohorts in these studies included patients who were acutely infected, whereas our study shows that HLA-DR downregulation can be prolonged months after infection. Parackova and colleagues found that monocyte HLA-DR in COVID-19 patients began to recover four weeks into hospital admission^30^. Thus, expression of HLA-DR on monocytes may indicate recovery, as the LR COVID PF cohort appeared to have low HLA-DR expression more than 1-month post-infection, while patients with ER COVID PF maintained or recovered HLA-DR expression.

Peripheral blood samples from patients with IPF also display decreased monocyte abundance and expression of MHC-II molecules on monocytes relative to age-matched controls. Monocyte depletion has been suggested to contribute to pathogenesis of IPF^53^, however more recent works have reported that higher monocyte abundance is correlated with poor outcomes in IPF^24,54–56^. Here, we recruited a clinically stable, outpatient IPF population, therefore, it may not reflect shifts in circulating monocyte populations during acute exacerbations or active progression of IPF. Notably, the present study indicates that HLA-DR expression is decreased exclusively on CD16+ monocytes in IPF, while conversely CD14+ and intermediate monocytes are the key populations affected in LR COVID PF. It is possible that our results in IPF indicate immunoparesis, while in COVID PF, enhanced migration of monocytes to the lung during COVID PF may evoke emergency myelopoiesis and eventually promote a state of exhaustion. Whether the observed decrease in HLA-DR+ monocytes in IPF and LR COVID PF arise from the same mechanism remains unknown, but further review of monocyte dysfunction in pulmonary fibrosis will require careful consideration of which monocyte subpopulations are affected.

Severe COVID-19 infection has been associated with an increase in CD8+ T cell activation^29,57^ as well as suppression of naïve CD4 T cells^8,58–61^. We observe evidence of each of these in T cell phenotypes in our LR COVID PF cohort. Compared to ER COVID PF patients, controls, and IPF patients, LR COVID PF patients had fewer naïve CD4+ T cells, and their CD8+ T cells expressed significantly greater levels of activation markers (HLA-DR and CD38). We therefore posit that the T cell response in LR COVID PF patients is polarized toward an effector or memory phenotype rather than naïve state, similar to what Su and colleagues described in post-acute sequelae of COVID-19 (PASC)^60^. This chronic inflammatory state may lead to senescence of CD8+ T cells, a state common in IPF^39–41^. Although our study did not observe gene expression changes suggestive of T cell senescence in the LR COVID PF cohort at the measured time point, McGroder and colleagues found that shortened telomere length, a defining characteristic of cellular senescence and feature associated with worse survival in IPF^62^, is an independent risk factor for developing fibrotic-like radiographic abnormalities after severe COVID-19^18^.

A limitation of the present study is that we restricted the age of our COVID PF study cohort (Mean (SD) age = 58±9 years old, Table E1). While this prevented age-associated changes from becoming a co-variant, it was necessary to properly compare our cohort to IPF patients. Similarly, our COVID PF cohort was majority male (13/16 patients) and sample size was limited. Further studies with a larger clinical cohort comprising young and old COVID PF patients as well as identifying immune signature changes in the lung are necessary to validate whether our findings apply to a broader population in the circulation and tissue.

Post-COVID fibrosis is an emerging cause of restrictive lung disease, and longitudinal studies are needed to evaluate the disease course in these patients. Our data imply the peripheral immune response of LR COVID PF is distinct from IPF. Our results suggest that in LR COVID PF patients, monocytes are either systemically depleted or alternatively recruited to the lung or other tissues. Studies confirm increased infiltration of monocyte-derived macrophages in the bronchoalveolar lavage fluid of severe COVID-19 patients^63^ and lungs of fatal COVID-19 ^64^. This observation presents the opportunity to investigate inhibiting monocyte recruitment as a method to improve recovery from COVID PF. Taken together, we propose that relative monocyte abundance may be a useful and simple prognostic indicator for determining whether long-haul COVID patients will resolve or have persistent pulmonary complications early in their disease course.

## Data Availability

All data produced in the present study are available upon reasonable request to the authors. Please email corresponding authors for supplemental tables and figures.

## Acknowledgments

The authors thank the UVA Genome Analysis and Technology Core for their technical expertise in single-cell RNA sequencing and the funding that supported this work: NHLBI 5K23HL143135-04, University of Virginia Engineering in Medicine Seed Fund, Global Infectious Diseases Institute COVID-19 Rapid Response (CAB), UVA Robert R. Wagner Fellowship (GCB), R21 AI160334 and U01 AI125056 (JAW).

## References

1. Logue, J. K. et al. Sequelae in Adults at 6 Months After COVID-19 Infection. JAMA Netw. Open 4, e210830 (2021).

2. Nalbandian, A. et al. Post-acute COVID-19 syndrome. Nat. Med. 27, 601–615 (2021).

3. Han, X. et al. Six-month Follow-up Chest CT Findings after Severe COVID-19 Pneumonia. Radiology 299, E177–E186 (2021).

4. Zhao, Y. et al. Follow-up study of the pulmonary function and related physiological characteristics of COVID-19 survivors three months after recovery. EClinicalMedicine 25, 100463 (2020).

5. Wild, J. M. et al. Understanding the burden of interstitial lung disease post-COVID-19: the UK Interstitial Lung Disease-Long COVID Study (UKILD-Long COVID). BMJ Open Respir. Res. 8, e001049 (2021).

6. Fabbri, L. et al. Post-viral parenchymal lung disease following COVID-19 and viral pneumonitis hospitalisation: A systematic review and meta-analysis. 2021.03.15.21253593 https://www.medrxiv.org/content/10.1101/2021.03.15.21253593v2 (2021) doi:10.1101/2021.03.15.21253593.

7. Raghu, G. et al. Idiopathic Pulmonary Fibrosis (an Update) and Progressive Pulmonary Fibrosis in Adults: An Official ATS/ERS/JRS/ALAT Clinical Practice Guideline. Am. J. Respir. Crit. Care Med. 205, e18–e47 (2022).

8. Zhao, Y. et al. Abnormal immunity of non-survivors with COVID-19: predictors for mortality. Infect. Dis. Poverty 9, 108 (2020).

9. Moderbacher, C. R. et al. Antigen-Specific Adaptive Immunity to SARS-CoV-2 in Acute COVID-19 and Associations with Age and Disease Severity. Cell 183, 996-1012.e19 (2020).

10. Zhou, X. & Ye, Q. Cellular Immune Response to COVID-19 and Potential Immune Modulators. Front. Immunol. 12, (2021).

11. Aghbash, P. S., Eslami, N., Shamekh, A., Entezari-Maleki, T. & Baghi, H. B. SARS-CoV-2 infection: The role of PD-1/PD-L1 and CTLA-4 axis. Life Sci. 270, 119124 (2021).

12. Giamarellos-Bourboulis, E. J. et al. Complex Immune Dysregulation in COVID-19 Patients with Severe Respiratory Failure. Cell Host Microbe 27, 992-1000.e3 (2020).

13. Torres Acosta, M. A. & Singer, B. D. Pathogenesis of COVID-19-induced ARDS: implications for an ageing population. Eur. Respir. J. 56, 2002049 (2020).

14. Shi, Y. et al. COVID-19 infection: the perspectives on immune responses. Cell Death Differ. 27, 1451–1454 (2020).

15. Sarkesh, A. et al. Extrapulmonary Clinical Manifestations in COVID-19 Patients. Am. J. Trop. Med. Hyg. 103, 1783–1796 (2020).

16. Cheon, I. et al. Immune signatures underlying post-acute COVID-19 lung sequelae. Sci. Immunol. 0, eabk1741.

17. Cocconcelli, E. et al. Characteristics and Prognostic Factors of Pulmonary Fibrosis After COVID-19 Pneumonia. Front. Med. 8, (2022).

18. McGroder, C. F. et al. Pulmonary fibrosis 4 months after COVID-19 is associated with severity of illness and blood leucocyte telomere length. Thorax 76, 1242–1245 (2021).

19. Vijayakumar, B. et al. Immuno-proteomic profiling reveals aberrant immune cell regulation in the airways of individuals with ongoing post-COVID-19 respiratory disease. Immunity 55, 542-556.e5 (2022).

20. Narasimhan, H., Wu, Y., Goplen, N. P. & Sun, J. Immune determinants of chronic sequelae after respiratory viral infection. Sci. Immunol. 7, eabm7996 (2022).

21. Herazo-Maya, J. D. et al. Validating a 52-gene risk profile for outcome prediction in Idiopathic Pulmonary Fibrosis: an international multicentre cohort study. Lancet Respir. Med. 5, 857–868 (2017).

22. O’Dwyer, D. N. et al. The peripheral blood proteome signature of idiopathic pulmonary fibrosis is distinct from normal and is associated with novel immunological processes. Sci. Rep. 7, 46560 (2017).

23. Unterman, A. et al. Single Cell RNA Sequencing of Peripheral Blood Mononuclear Cells in Idiopathic Pulmonary Fibrosis Reveals the Cellular Origin of the Outcome Predictive 52-Gene Signature. in D13. ILD PROGNOSIS AND BIOMARKERS II A6211–A6211 (American Thoracic Society, 2020). doi:10.1164/ajrccm-conference.2020.201.1_MeetingAbstracts.A6211.

24. Scott, M. K. D. et al. Increased monocyte count as a cellular biomarker for poor outcomes in fibrotic diseases: a retrospective, multicentre cohort study. Lancet Respir. Med. 7, 497–508 (2019).

25. Huang, Y. et al. A functional genomic model for predicting prognosis in idiopathic pulmonary fibrosis. BMC Pulm. Med. 15, 147 (2015).

26. Ramani, C. et al. Post-ICU COVID-19 Outcomes: A Case Series. Chest 159, 215– 218 (2021).

27. Augustin, M. et al. Post-COVID syndrome in non-hospitalised patients with COVID-19: a longitudinal prospective cohort study. Lancet Reg. Health – Eur. 6, (2021).

28. Bell, M. L. et al. Post-acute sequelae of COVID-19 in a non-hospitalized cohort: Results from the Arizona CoVHORT. PLOS ONE 16, e0254347 (2021).

29. Unterman, A. et al. Single-cell multi-omics reveals dyssynchrony of the innate and adaptive immune system in progressive COVID-19. Nat. Commun. 13, 440 (2022).

30. Parackova, Z. et al. Disharmonic Inflammatory Signatures in COVID-19: Augmented Neutrophils’ but Impaired Monocytes’ and Dendritic Cells’ Responsiveness. Cells 9, 2206 (2020).

31. Silvin, A. et al. Elevated Calprotectin and Abnormal Myeloid Cell Subsets Discriminate Severe from Mild COVID-19. Cell 182, 1401-1418.e18 (2020).

32. Wilk, A. J. et al. A single-cell atlas of the peripheral immune response in patients with severe COVID-19. Nat. Med. 26, 1070–1076 (2020).

33. Arunachalam, P. S. et al. Systems biological assessment of immunity to mild versus severe COVID-19 infection in humans. Science 369, 1210–1220 (2020).

34. Finak, G. et al. MAST: a flexible statistical framework for assessing transcriptional changes and characterizing heterogeneity in single-cell RNA sequencing data. Genome Biol. 16, 278 (2015).

35. Subramanian, A. et al. Gene set enrichment analysis: A knowledge-based approach for interpreting genome-wide expression profiles. Proc. Natl. Acad. Sci. 102, 15545– 15550 (2005).

36. Liberzon, A. et al. The Molecular Signatures Database (MSigDB) hallmark gene set collection. Cell Syst. 1, 417–425 (2015).

37. Lee, J. S. et al. Immunophenotyping of COVID-19 and influenza highlights the role of type I interferons in development of severe COVID-19. Sci. Immunol. 5, eabd1554 (2020).

38. Zhang, Q. et al. Inborn errors of type I IFN immunity in patients with life-threatening COVID-19. Science 370, eabd4570 (2020).

39. Bonham, C. A. et al. T cell Co-Stimulatory molecules ICOS and CD28 stratify idiopathic pulmonary fibrosis survival. Respir. Med. X 1, (2019).

40. Habiel, D. M. et al. Characterization of CD28null T cells in idiopathic pulmonary fibrosis. Mucosal Immunol. 12, 212–222 (2019).

41. Moore, C. et al. Resequencing Study Confirms That Host Defense and Cell Senescence Gene Variants Contribute to the Risk of Idiopathic Pulmonary Fibrosis. Am. J. Respir. Crit. Care Med. 200, 199–208 (2019).

42. Shen, Z. et al. miR-200b regulates cellular senescence and inflammatory responses by targeting ZEB2 in pulmonary emphysema. Artif. Cells Nanomedicine Biotechnol. 48, 656–663 (2020).

43. Crawford, A. et al. Molecular and transcriptional basis of CD4+ T cell dysfunction during chronic infection. Immunity 40, 289–302 (2014).

44. Effros, R. B. Loss of CD28 expression on T lymphocytes: a marker of replicative senescence. Dev. Comp. Immunol. 21, 471–478 (1997).

45. Schulte-Schrepping, J. et al. Severe COVID-19 Is Marked by a Dysregulated Myeloid Cell Compartment. Cell 182, 1419-1440.e23 (2020).

46. Handy, J. M. et al. HLA-DR expression and differential trafficking of monocyte subsets following low to intermediate risk surgery*. Anaesthesia 65, 27–35 (2010).

47. Townsend, L. et al. Severe COVID-19 is characterised by inflammation and immature myeloid cells early in disease progression. Heliyon 8, e09230 (2022).

48. Interleukin 10 (IL-10) and viral IL-10 strongly reduce antigen-specific human T cell proliferation by diminishing the antigen-presenting capacity of monocytes via downregulation of class II major histocompatibility complex expression. J. Exp. Med. 174, 915–924 (1991).

49. Greco, M. et al. Human Leukocyte Antigen-DR Isotype Expression in Monocytes and T Cells Interferon-Gamma Release Assay in Septic Patients and Correlation With Clinical Outcome. J. Clin. Med. Res. 13, 293–303 (2021).

50. Lukaszewicz, A.-C. et al. Monocytic HLA-DR expression in intensive care patients: interest for prognosis and secondary infection prediction. Crit. Care Med. 37, 2746– 2752 (2009).

51. Payen, D. et al. A Longitudinal Study of Immune Cells in Severe COVID-19 Patients. Front. Immunol. 11, (2020).

52. Hammad, R. et al. Utility of Monocyte Expression of HLA-DR versus T Lymphocyte Frequency in the Assessment of COVID-19 Outcome. Int. J. Gen. Med. 15, 5073– 5087 (2022).

53. Greiffo, F., Fernandez, I., Frankenberger, M., Behr, J. & Eickelberg, O. Circulating monocytes from interstitial lung disease patients show an activated phenotype. Eur. Respir. J. 48, (2016).

54. Achaiah, A. et al. Monocyte and neutrophil levels are potentially linked to progression to IPF for patients with indeterminate UIP CT pattern. BMJ Open Respir. Res. 8, e000899 (2021).

55. Karampitsakos, T. et al. Increased monocyte count and red cell distribution width as prognostic biomarkers in patients with Idiopathic Pulmonary Fibrosis. Respir. Res. 22, 140 (2021).

56. Kreuter, M. et al. Monocyte Count as a Prognostic Biomarker in Patients with Idiopathic Pulmonary Fibrosis. Am. J. Respir. Crit. Care Med. 204, 74–81 (2021).

57. Du, J. et al. Persistent High Percentage of HLA-DR+CD38high CD8+ T Cells Associated With Immune Disorder and Disease Severity of COVID-19. Front. Immunol. 12, 735125 (2021).

58. Phetsouphanh, C. et al. Immunological dysfunction persists for 8 months following initial mild-to-moderate SARS-CoV-2 infection. Nat. Immunol. 23, 210–216 (2022).

59. Diao, B. et al. Reduction and Functional Exhaustion of T Cells in Patients With Coronavirus Disease 2019 (COVID-19). Front. Immunol. 11, 827 (2020).

60. Su, Y. et al. Multiple early factors anticipate post-acute COVID-19 sequelae. Cell 185, 881-895.e20 (2022).

61. de Candia, P., Prattichizzo, F., Garavelli, S. & Matarese, G. T Cells: Warriors of SARS-CoV-2 Infection. Trends Immunol. 42, 18–30 (2021).

62. Stuart, T. et al. Comprehensive Integration of Single-Cell Data. Cell 177, 1888-1902.e21 (2019).

63. Liao, M. et al. Single-cell landscape of bronchoalveolar immune cells in patients with COVID-19. Nat. Med. 26, 842–844 (2020).

64. Melms, J. C. et al. A molecular single-cell lung atlas of lethal COVID-19. Nature 595, 114–119 (2021).

