## Supplemental Methods for "Reduction in circulating monocytes correlates with persistent post-COVID pulmonary fibrosis in multi-omic comparison of long-haul COVID and IPF"

*Sample Collection*
Patients who had IPF or had recovered from COVID-19 were recruited through the University of Virginia (UVA) Pulmonary and Post-COVID clinic respectively. Peripheral blood mononuclear cells (PBMCs) were isolated from venous blood (Post-COVID and IPF: K2EDTA BD Vacutainer®) by density gradient centrifugation and cryopreserved for later analysis (FBS + 10% DMSO). PBMC collection and single cell RNA sequencing for control samples were prepared at the Mayo Clinic similarly to samples detailed by Cheon et al. [1]

*Ethics Statement*
All studies performed at UVA were approved by the UVA Human Investigations Committee. Studies performed at the Mayo Clinic were approved by Mayo Clinic Institutional Review Boards (protocol ID 20-004911). All subjects provided written informed consent.

*Single Cell RNA Sequencing Sample Preparation*
The generation of single cell indexed libraries, from the thawed PBMC, was performed by the School of Medicine Genome Analysis and Technology Core, RRID:SCR_018883, using the 10X Genomics chromium controller platform and the Chromium Single Cell 5′ Library & Gel Bead Kit v1.1 reagent. Briefly, around 10,000 cells were targeted per sample and loaded onto each well of a Chromium Single Cell G Chip to generate single cell emulsions primed for reverse transcription. After breaking the emulsion, the single cell specific barcoded DNAs were subjected to cDNA amplification and QC on the Agilent 4200 TapeStation Instrument, using the Agilent D5000 kit. Each sample cDNA was used to prepare indexed libraries that were pooled prior to sequencing. A QC run was performed on the Illumina Miseq using the nano 300Cycle kit (1.4 Million reads/run), to estimate the number of targeted cells per sample using the Cellranger 3.0.2 function. The cell estimate enabled the core to re-balance the pooled sample prior to deep sequencing onto the NextSeq 2000, using the P3-100 cycle kit. After run completion, the Binary base call (bcl) files were converted to fastq format using the Illumina bcl2fastq2 software, and data transferred to the Bioinformatics core for further analysis. To accommodate batch effects, all samples were prepared with different treatments (ER COVID PF, LR COVID PF, and IPF). This technique was successful the vast majority of our data with the exception of the 3 non-diseased controls prepared at the Mayo Clinic and 3 of the 18 single-cell RNA seq samples prepared at UVA where the complementary samples failed quality control for that batch.

*Single Cell RNA Sequencing Data Cleaning and Integration*
Raw fastq files from all 21 samples were aligned and quantified using a human reference genome. Following the suggested pipeline for quality control in the Seurat v.4.0.3 package, sc-RNAseq data were filtered for dead cells, doublets, and red blood cells by excluding cells with greater than 5% mitochondrial genes and less than 500, but no more than 2500 genes. Samples underwent normalization, scaling, integration using anchors, dimensional reduction, and further downstream analysis using the standard

Seurat workflow with the Seurat v.4.0.3 package. PCs were visualized using an elbow plot to select dimensionality of the 21-sample integrated dataset. From this quantitative approach, we determined to implement 17 dimensions as our input for the RunUMAP and FindNeighbors clustering parameters and set the resolution to 0.5 for our initial clustering. Nineteen cell clusters were identified initially. Erythrocyte clusters were removed, and two CD4+ T effector cell clusters were merged, yielding 16 distinct immune cell subpopulations (69,868 cells). Dot plots depicting the defining markers for each cluster were constructed using default settings in the DotPlot function.

*Single Cell RNA Sequencing Cell Abundance Quantification*
Labels denoting each condition were appended to each cluster ID and counted using the Idents function in Seurat. These values were then exported to Excel where relative cell abundance to the total cell population were calculated ( (number of cells in cluster x / sum of cells from all clusters)*100). Total cell population was replaced with all CD4 T cells or all CD8 T cells when applicable for T cell subset relative abundances. All bar charts representing relative cell abundance were constructed and statistically analyzed in GraphPad Prism 9.4.1. To determine whether the time between initial COVID infection and sample collection (in months) was a potential cofounding variable for cell abundance Pearson or Spearman Correlations (depending on normality determined by Shapiro-Wilk Test) were performed.

*Differential Expressed Gene and Gene Set Enrichment Analyses*
Differentially expressed genes (DEGs) between conditions (non-diseased control, ER COVID PF, LR COVID PF, and IPF) were determined using the FindMarkers function in Seurat. Significance for the difference in gene expression between the two groups were tested using the default Wilcox rank sum test and adjusted with bonferroni correction using all genes to account for false discovery. Violin plots depicting DEGs were constructed using default settings in the VlnPlot function. To account for time between initial COVID infection and sample collection (in months) as a potential cofounding variable we performed a multiple linear regression analysis using the latent.vars argument in the FindMarkers function.

Differentially expressed genes for gene set enrichment analysis (GSEA) were determined by applying model-based analysis of single-cell transcriptomics (MAST) test and the clusterProfiler package to the whole gene expression profile of ER versus LR COVID PF as well as each LR COVID PF versus the non-diseased control group. The canonical pathways from the curated gene set (C2) provided by Molecular Signatures Database (MSigDB) were input as the gene list for GSEA. GSEA results were output as a table found in supplemental or visualized using the dotplot function from ggplot package.

Similarly, MAST DEG outputs were used to generate Volcano Plots using the EnhancedVolcano and tidyverse packages.

*Single Cell RNA Sequencing Re-clustering*

All CD4+ T cell populations were merged into one cluster and subset out for CD4+ T cell re-clustering. The subset object containing all CD4+ T cells was then re-processed for variable features using the vst selection method and setting nfeatures to 2000, followed by re-scaling in Seurat. For dimensional reduction, the dimension argument was set to 8 for the FindNeighbors function and the resolution to 0.75 for the FindClusters function. Nine populations were output from this analysis. Clusters 0 and 7 expressed similarly high levels of the naïve markers CCR7+ and SELL+ without expressing effector markers and therefore were combined into one Naïve CD4+ T cell population. Similarly, clusters 5 and 1 were combined as these clusters expressed lower levels of CCR7 and SELL while expressing similar amounts of the memory markers CD27, S100A4, and PASK. We identified a naïve subpopulation, an Early Activation subpopulation (denoted by high expression of naïve markers with moderate expression of activation markers, such as CD69+, suggesting these naïve cells were recently stimulated and in the early response of transitioning to an effector or memory function), Th1-like (TBX21+, CXCR3+), Th2-like (GATA3hi, CCR4+), Th17-like (RORC+, CCR6+), and Treg (FOXP3+, IL2RA+).

*Sample Multiplex Staining*

Briefly, PBMCs were spun down at 300 rcf for 5 minutes and washed in Hydration Buffer (Akoya). Cells were then resuspended in 1.6% PFA diluted in Hydration Buffer and fixed for 20 minutes at room temperature on a rotator. The sample was then spun down and washed in Hydration Buffer again as stated above. Once the supernatant was removed, fixed PBMCs were then seeded onto a poly-L-lysine coated coverslip and allowed to air dry for 10 minutes. Dried coverslips were washed in PBS two times, incubated with Staining Buffer (Akoya) for 20 minutes, and then placed in a humidity chamber for staining with the 13 antibody panel. All samples were stained for three hours at room temperature in the humidity chamber with CD45 (Catalog # 4150003), CD2 (Catalog # 4250005), CD19 (Catalog # 4350003), CD38 (Catalog # 4150007), CD11c (Catalog # 4350012), CD278 (Catalog # 4250013), CD8 (Catalog # 4150004), CD3 (Catalog # 4350008), CD69 (Catalog # 4250022), CD4 (Catalog # 4350010), Ki67 (Catalog # 4250019), CD279 (Catalog # 4250010), and HLA-DR (Catalog # 4250006) using Akoya manufacturer's instructions and blockers (Further information on antibodies in Table E2). Coverslips were then washed in Staining Buffer two times, followed by a post-staining fixation with 1.6% PFA in Storage Buffer (Catalogue # 232107) for 10 minutes at room temperature. Slides were washed three times in PBS and incubated in ice cold methanol for 5 minutes. Samples were washed three times with PBS and then prepared for a final fixation in fresh BS3 diluted in PBS for 20 minutes before being washed in PBS three times and stored at 4 degrees C in Storage Buffer until imaging.

*Multiplex Imaging and Processing*

Samples stained with the 13-antibody immune panel underwent multiplexed imaging using the spatial-omics platform PhenoCycler (Akoya) in combination with the BZ-X810 slide scanning microscope (Keyence). Further detail on automated imaging acquisition and fluidics exchange using the PhenoCycler is described by Goltsev et al. and Schürch et al.[2,3]. Raw TIFF images were stitched and processed using the PhenoCycler Processor which executes drift compensation, deconvolution, background subtraction, cycle alignment, and cell segmentation via a watershed cell segmentation algorithm. Data was then visualized and analyzed in MAV (Multiplex Analysis Viewer), a FIJI/ImageJ plugin.

Flow cytometry standard (FCS) files were generated from the processed data for each sample and imported to FCS Express 7 for further analysis.

*Quantification of Protein Expression and Cell Gating*

FCS files for each image region generated by the PhenoCycler were concatenated and analyzed in FCS Express 7. Gates to determine cell type and protein expression were tailored to each sample in a blinded fashion by experienced flow cytometrist. Samples were gated on DAPI and CD45 double positive cells to identify nucleated PBMCs. Gating strategy for determining major cell types (CD4+ T cell, CD8+ T cell, CD11c+ Monocytes, CD3-CD2-CD19-CD11c- Natural Killer-like cells) is shown in Figure E1. Gates to quantify percent of cells expressing HLA-DR, CD69, CD38, ICOS, PD-1, and Ki-67 were kept consistent when possible for each imaging batch, and known negative cell populations were used as an internal control, where possible. Signal and gating were confirmed by referencing processed images in MAV. All bar charts representing relative cell abundance and protein expression were constructed in GraphPad Prism 9.4.1.

*Statistical Tests*

Details regarding statistic test used for each analysis are included above or in the respective figure caption. All statistical tests were performed in R Studio 4.1.1 or GraphPad Prism 9.4.1 (Welch's t-test, Pearson correlations, one-way ANOVAs, and Kruskal-Wallis).

**References:**


1. Cheon, I. *et al.* Immune signatures underlying post-acute COVID-19 lung sequelae. *Sci. Immunol.* **0**, eabk1741.
2. Goltsev, Y. *et al.* Deep Profiling of Mouse Splenic Architecture with CODEX Multiplexed Imaging. *Cell* **174**, 968-981.e15 (2018).
3. Schürch, C. M. *et al.* Coordinated Cellular Neighborhoods Orchestrate Antitumoral Immunity at the Colorectal Cancer Invasive Front. *Cell* **182**, 1341-1359.e19 (2020).
